# A multi-ancestry polygenic risk score for body mass index predicts longitudinal weight change

**DOI:** 10.1101/2025.07.17.25331705

**Authors:** Tianyuan Lu, Lily N Stalter, Kate V Lauer, Bret M Hanlon, Wenmin Zhang, Luke M Funk

## Abstract

**Background:** Persistent weight gain directly leads to obesity. Identifying individuals at risk for future weight gain is challenging, partly because associations with traditional clinical risk factors may be biased by confounding and reverse causation. Polygenic risk scores (PRS) provide a stable, lifelong measure of genetic predisposition to obesity. However, existing PRS have not been evaluated for their association with longitudinal weight change in adulthood and often lack generalizability across diverse genetic ancestry groups.

**Methods:** We conducted ancestry-specific genome-wide association study meta-analyses of body mass index (BMI) in populations of European, African or African American, Admixed American, East Asian, and South Asian ancestries and developed ancestry-specific PRS. A multi-ancestry polygenic risk score (MAPRS) was trained using ancestry-specific PRS in a model selection dataset (N=39,685) from the NIH All of Us Research Program (AoU). We evaluated the MAPRS in an independent AoU model evaluation dataset (N=158,743) for BMI prediction and in a separate AoU test dataset (N=78,219) with repeated measurements over 1.5–2.5 years for weight change prediction. The outcomes included change in BMI and ≥10% or ≥5% total body weight (TBW) gain. We further examined the relationship between MAPRS and 12 clinical risk factors commonly comorbid with obesity in relation to weight change.

**Results:** The MAPRS captured 7.05% of the variance in measured BMI in the AoU model evaluation dataset and demonstrated improved generalizability across all non-European genetic ancestry groups. In the AoU test dataset, conditioned on baseline BMI at the second-to-last measurement, a one SD increase in MAPRS was associated with a 0.16 kg/m^2^ increase in future BMI (standard error=0.012 kg/m^2^; p-value=2.2×10^−39^), 1.27-fold increased odds of experiencing ≥10% TBW gain (95% CI: 1.24-1.31; p-value=1.4×10^−55^), and 1.15-fold increased odds of experiencing ≥5% TBW gain (95% CI: 1.13-1.18; p-value=2.8×10^−39^). These associations were observed across all genetic ancestry groups and remained highly consistent after adjustment for any clinical risk factor. In contrast, most clinical risk factors demonstrated inconsistent or weaker associations with weight change outcomes.

**Conclusions:** The MAPRS for BMI is a robust and generalizable risk factor for weight gain. Its potential utility for early risk stratification and targeted prevention warrants further investigation.

## Background

Obesity is a pressing global public health challenge, contributing significantly to the development of numerous chronic conditions and imposing a growing burden on healthcare systems and individual well-being^1–3^. Globally, 2.5 billion adults met the body mass index (BMI) criterion for overweight (BMI > 25 kg/m^2^) in 2022, and approximately 890 million met the criterion for obesity (BMI > 30 kg/m^2^), with prevalence continuing to rise^4^. Clinically significant increases in body weight, such as gains of at least 5% in total body weight (TBW), are strongly associated with the onset of obesity-related comorbidities, including type 2 diabetes, cardiovascular disease, and certain cancers^5–7^.

Early identification of individuals at elevated risk of future weight gain is critical, as it may facilitate preventive interventions, such as lifestyle modifications, nutritional guidance, behavioral counseling, or pharmacotherapy, before patients develop severe obesity (BMI > 35 kg/m^2^) and/or obesity-related comorbidities. However, this remains a major clinical challenge, because the relationship between many clinical conditions and obesity can be confounded or subject to reverse causation, where the clinical conditions can develop as consequences of prior weight gain. This complicates the interpretation of observed associations and limits their utility as predictive markers, highlighting the need for more reliable and temporally stable indicators of future weight gain risk.

Genetic factors have been recognized as important contributors to obesity^8^. Measures of obesity, such as BMI, are highly polygenic traits, influenced by the cumulative effects of genetic variants across the genome^9–11^. These variants impact a wide range of biological pathways, including those involved in appetite regulation, energy expenditure, adipose tissue biology, and neuroendocrine function^9–11^. Polygenic risk scores (PRS), which quantify the aggregated the effects of these variants, have emerged as powerful tools for capturing inherited predisposition to various complex traits^12–17^. PRS for BMI have demonstrated strong correlations with BMI and other obesity-related traits in cross-sectional analyses^12,18–20^. However, it remains unclear whether these scores can predict weight changes over time, which is a more dynamic and clinically relevant outcome. Such longitudinal prediction is particularly important for individuals who do not yet meet the criteria for obesity but may be on a trajectory toward obesity and for whom early intervention can be most impactful. Furthermore, most existing PRS are based on genome-wide association studies (GWAS) conducted primarily in populations of European ancestry, which limits their generalizability and predictive accuracy across diverse populations^21^.

To address these limitations, in this study, we developed a multi-ancestry polygenic risk score (MAPRS) for BMI by integrating existing GWAS data from five continental genetic ancestry groups. Leveraging the National Institutes of Health (NIH) All of Us Research Program^22^ (AoU; version 8), we evaluated the MAPRS in relation to both measured BMI and prospective weight change over a follow-up period of 1.5–2.5 years in a multi-ancestral population. We further assessed the relative strength and stability of genetic risk compared to 12 clinical risk factors commonly comorbid with obesity in predicting weight gain. This study provides key insights into the utility of an MAPRS as a robust and generalizable risk factor for future weight gain and offers a foundation for its potential use in preventive healthcare and early intervention strategies.

## Methods

### Ancestry-specific meta-analyses of genome-wide association studies

To maximize the statistical power for detecting genetic associations with BMI, we conducted ancestry-specific GWAS meta-analyses for European, African and African American (collectively referred to as African hereafter), East Asian, and South Asian ancestry populations across several large cohorts. The GWAS included in this study are summarized in Supplementary Table 1. In all cohorts, inverse normal-transformed BMI values were used as the outcome in GWAS. For individuals of European ancestry, we included data from the U.S. Veterans Health Administration (VA) Million Veteran Program^23^ (MVP; up to 424,221 individuals), the Genetic Investigation of ANthropometric Traits^10^ (GIANT) consortium (up to 322,154 individuals), UK Biobank^24^ (up to 419,163 individuals), and FinnGen^25^ (up to 500,348 individuals). For African ancestry, we included MVP^23^ (up to 118,993 individuals), UK Biobank^24^ (up to 6,545 individuals), the Uganda Genome Resource^26^ (up to 6,197 individuals), the Durban Diabetes Study^26^ (up to 1,114 individuals), the Diabetes Case Control Study^26^ (up to 1,478 individuals), and the Africa America Diabetes Mellitus Study^26^ (up to 5,187 individuals). For East Asian ancestry, we included MVP^23^ (up to 6,384 individuals), UK Biobank^24^ (up to 2,693 individuals), BioBank Japan^27^ (up to 163,835 individuals), and Taiwan Biobank^28^ (up to 92,615 individuals). For South Asian ancestry, we included UK Biobank^24^ (up to 8,646 individuals) and East London Genes & Health^29^ (up to 34,408 individuals). Within each genetic ancestry group, we performed inverse variance-weighted meta-analysis of the effect size estimates using METAL^30^. Additionally, we obtained Admixed American ancestry-specific GWAS summary statistics from MVP^23^ (up to 57,793 individuals). There was no known sample overlap between these cohorts and AoU.

### National Institutes of Health All of Us Research Program

The AoU is a large, diverse cohort based in the United States launched in 2018^22^. Data in AoU (version 8) were collected through electronic health records, self-reported surveys, physical measurements and biospecimen collection. For this study, we included participants aged 18 to 70 years at the time of measurement, between 2008 and 2023, who did not have a diagnosis of any type of cancer, or a history of bariatric surgery based on International Classifications of Diseases (ICD) and Systematized Nomenclature of Medicine codes in electronic health records (Supplementary Table 2). To ensure data quality, we applied a stringent phenotype quality control protocol based on the methods described by Cheng et al^31^. Specifically, we excluded biologically implausible height (< 111.8 cm [44 inches] or > 228.6 cm [90 inches]) and weight (< 24.9 kg [55 pounds] or > 453.6 kg [1,000 pounds]) values. Additional outlier detection was performed based on individual-level distributions. Weight values were considered inaccurate if: (1) the range exceeded 22.7 kg and the absolute deviation from the mean was > 70% of the range, or (2) the standard deviation exceeded 20% of the mean and the absolute deviation from the mean exceeded the standard deviation. Height values were considered inaccurate if: (1) the absolute deviation from the mean exceeded the standard deviation, and (2) the standard deviation was > 2.5% of the mean. When multiple valid height measurements were available, the most recent value was used to calculate BMI.

The All of Us Genome Centers generated whole-genome sequencing data from DNA extracted from blood samples using the Illumina NovaSeq 6000 platform, following standardized protocols for quality control and data harmonization^22^. Individuals whose DNA samples failed sample outlier quality control were excluded from analysis. We excluded individuals with a kinship coefficient >0.1 to reduce potential bias from relatedness. DNA samples that failed sample outlier quality control were excluded. Genetic ancestry in AoU was assigned by projecting samples into principal component space derived from the Human Genome Diversity Project^32^ and 1000 Genomes Project^33^ reference data and applying a random forest classifier trained on known genetic ancestry labels, as described previously^22^.

For downstream analyses, individuals with only one valid BMI measurement were randomly split into a model development dataset (20%; Supplementary Table 3) and a model evaluation dataset (80%; Supplementary Table 3), excluding those with a BMI value < 12 kg/m^2^ or > 70 kg/m^2^. A separate test dataset was constructed from individuals using the two most recent valid BMI values measured approximately two years apart (1.5–2.5 years; Supplementary Table 4), excluding those with a baseline BMI value < 12 kg/m^2^ or > 70 kg/m^2^ at the second-to-last measurement. Participants with a recorded pregnancy with one year of BMI measurement were also excluded (Supplementary Table 2).

### Development and validation of a multi-ancestry polygenic risk score for body mass index

We followed an established approach^14,34^ to construct an MAPRS for BMI. We first generated ancestry-specific PRS using results of the ancestry-specific GWAS meta-analyses. For each GWAS, we retained variants present in AoU with a population-specific allele frequency > 1% or allele count > 100. We performed linkage disequilibrium (LD) clumping using PLINK^35^ (version 1.9 beta) with the corresponding ancestry-matched LD reference panel from the 1000 Genomes Project^33^. LD clumping was performed with a window size of 10 megabases (clump_kb = 10,000), an LD R^2^ threshold of 0.01 (clump_r2 = 0.01), and a p-value threshold of 1.0×10^−5^ to retain suggestively significant variants. The resulting variants and their corresponding effect size estimates were then used to compute ancestry-specific PRS in AoU. We combined the five ancestry-specific PRS by fitting a multivariable linear regression model in the AoU model development dataset of the form:

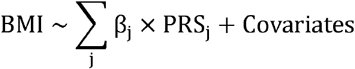

which included j ∈ {European, African, Admixed American, East Asian, South Asian}, and age at measurement, biological sex (male or female), and the first 10 genetic principal components as covariates. The estimated coefficients 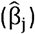 for each ancestry-specific PRS served as weights for combining them into the final MAPRS^14,34^.

We evaluated the predictive performance of the MAPRS in the independent AoU model evaluation dataset, both overall and stratified by genetic ancestry groups. Performance of the MAPRS was assessed using the increment in the proportion of variance explained compared to a baseline model, defined as:

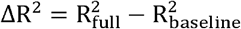

Where 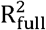 represents the proportion of variance in BMI explained by a linear regression model baseline including the MAPRS, age at measurement, sex, and the first 10 genetic principal components as predictors, and 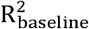 represents the proportion of variance in BMI explained by a linear regression model including only age at measurement, sex, and the first 10 genetic principal components as predictors.

The MAPRS was benchmarked against (1) each of the five ancestry-specific PRS, and (2) three previously developed PRS for BMI based on European ancestry GWAS, using direct LD clumping and thresholding (PGS000717^19^), LDpred^36^ (PGS000027^12^), and penalized regression (PGS001228^20^), respectively. Variant weights in the previously developed PRS were obtained from the Polygenic Score Catalog^37^. We calculated Δ R^2^as defined above for each of these scores in the AoU model evaluation dataset.

### Evaluation of polygenic risk score as a risk factor for weight gain

We evaluated the association between the MAPRS and longitudinal weight change over 1.5-2.5 years in the AoU test dataset. Three weight change outcomes were considered:

1. Change in BMI, calculated as the difference between the most recent and baseline (i.e., second-to-last) BMI measurements. This outcome captures weight change adjusted for height.
2. ≥10% total body weight (TBW) gain, defined as a binary outcome indicating whether an individual’s weight increased by at least 10% between the two time points, irrespective of height.
3. ≥5% TBW gain, defined similarly as a binary outcome for at least 5% weight increase.

We assessed the association between the MAPRS and change in BMI using linear regression, adjusted for baseline age (i.e., age at the second-to-last measurement), sex, and baseline BMI (i.e., BMI at the second-to-last measurement). We assessed the association between the MAPRS and ≥10% or ≥5% TBW gain using logistic regression, adjusted for the same covariates. These analyses were performed both in the overall AoU test dataset, where genetic ancestry was included as an additional covariate, and separately within each genetic ancestry group. Furthermore, we conducted sex-stratified analyses to evaluate whether these associations were consistent between males and females. Prior to each analysis, the MAPRS was standardized to have zero mean and unit variance within the corresponding subset.

### Assessment of the relationship between polygenic risk score and clinical risk factors in association with weight change

We examined the relationship between the MAPRS and 12 clinical risk factors commonly comorbid with obesity. These risk factors included hypertension, hyperlipidemia, type 2 diabetes, obstructive sleep apnea, gastroesophageal reflux disease (GERD), cardiovascular disease (CVD), osteoarthritis, chronic pain, depression, anxiety, non-alcoholic fatty liver disease (NAFLD), and cholelithiasis. All clinical risk factors were defined using ICD codes in electronic health records prior to the most recent BMI measurement (Supplementary Table 2).

We first assessed the association between the MAPRS and each of these clinical risk factors using logistic regression, with the MAPRS as the independent variable, adjusting for baseline age and sex. To investigate whether the MAPRS may act partially through these clinical risk factors, we then repeated the MAPRS-weight change outcome association analyses. These included change in BMI modeled using linear regression, and ≥10% and ≥5% TBW gain modeled using logistic regression, with additional adjustment for the clinical risk factors. Specifically, we added each clinical risk factor individually to models that included the MAPRS, baseline age, sex, and baseline BMI, and, in separate analyses, added all 12 clinical risk factors simultaneously. Attenuation of the association between the MAPRS and weight change outcomes after adjustment would suggest potential mediation or confounding by the corresponding risk factors. These analyses were performed both in the overall AoU test dataset, where genetic ancestry was included as an additional covariate, and separately within each genetic ancestry group. Sex-stratified analyses were also conducted. Prior to each analysis, the MAPRS was standardized to have zero mean and unit variance within the corresponding subset.

Finally, we assessed the magnitude of association between each clinical risk factor and each weight change outcome, adjusting for baseline age, sex, baseline BMI, and genetic ancestry. These associations were then compared to those between the MAPRS and the weight change outcomes to evaluate the relative contributions of genetic and clinical risk factors within the AoU test dataset. An association was considered significant with a p-value < 1.3×10^−3^ (= 0.05/13/3), corresponding to a Bonferroni correction accounting for 13 genetic or clinical risk factors and three weight change outcomes.

## Results

### Study overview

An overview of the study design is presented in Figure 1. Meta-analyses of GWAS were performed based on cohorts with no known overlap with AoU. Development and validation of the MAPRS were conducted based on the AoU model development dataset (N = 39,685) and the AoU model evaluation dataset (N = 158,743; Supplementary Table 3), respectively. The AoU model development dataset had a mean age of 46.5 years (standard deviation, SD = 15.3 years) and a mean BMI of 29.3 kg/m^2^ (SD = 7.0 kg/m^2^), while the AoU model evaluation dataset had a mean age of 46.6 years (SD = 15.3 years) and a mean BMI of 29.4 kg/m^2^ (SD = 7.0 kg/m^2^). The AoU test dataset (N = 78,219) was used to assess the association between the MAPRS and weight change outcomes. Compared to the model development and evaluation datasets, individuals in the AoU test dataset were older (mean baseline age = 49.6 years at the second-to-last measurement; SD = 14.0 years) and had higher BMI (mean BMI = 30.0 kg/m^2^; SD = 7.0 kg/m^2^ at both baseline and the most recent measurement), with this pattern consistent across all genetic ancestry groups (Supplementary Table 4). The mean BMI change was minimal, at - 0.028 kg/m^2^ (SD = 2.7 kg/m^2^). Overall, 7,618 (9.7%) participants experienced ≥10% TBW gain, and 18,533 (23.4%) had ≥5% TBW gain between the two most recent measurements.

**Figure 1.**
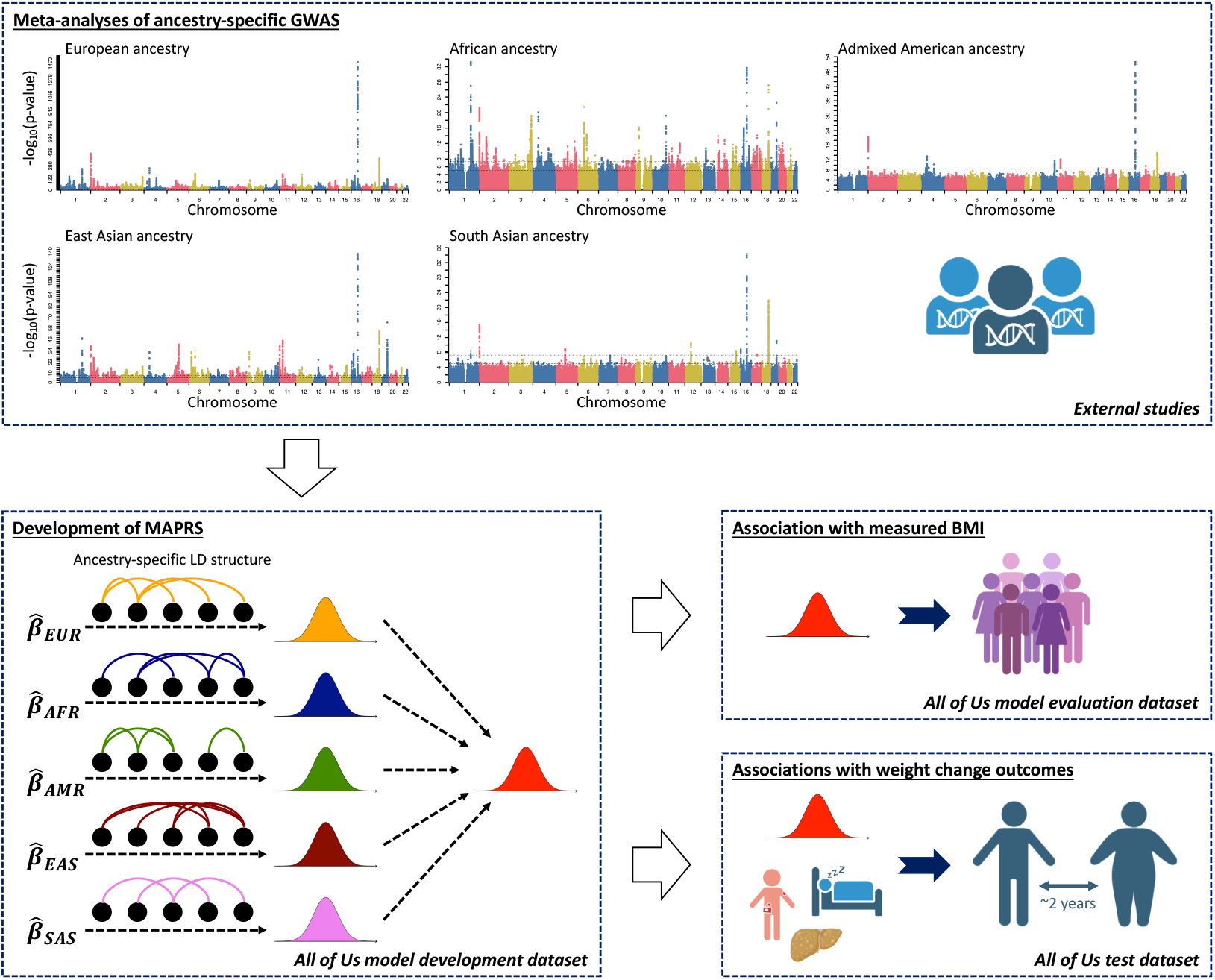
Overview of study design. Meta-analyses of ancestry-specific GWAS were conducted using summary statistics from large-scale cohorts. The AoU dataset was divided into a model development dataset, a model evaluation dataset, and a test dataset (Methods). The model development dataset (N = 39,685) and the model evaluation dataset (N = 158,743) comprised 20% and 80%, respectively, of the participants with one valid BMI measurement, while the test dataset (N = 78,219) included participants with at least two valid BMI measurements. The MAPRS was trained in the model development dataset by combining ancestry-specific polygenic risk scores for European (EUR), African or African American (AFR), Admixed American (AMR), East Asian (EAS), and South Asian (SAS) ancestry groups. Its performance in predicting BMI was assessed in the model evaluation dataset. Associations between the MAPRS, 12 clinical risk factors, and weight change outcomes were evaluated in the test dataset.

Across all datasets, East Asian and South Asian ancestry participants were younger, had lower mean BMI, and showed lower prevalence of most clinical risk factors compared to other ancestry groups (Supplementary Tables 3 and 4). African and Admixed American ancestry participants were also younger than European ancestry participants but had higher mean BMI (Supplementary Tables 3 and 4). The proportion of individuals with ≥10% TBW gain was highest among Admixed American ancestry participants (11.0%), followed by African (10.4%), European (9.1%), South Asian (8.3%), and East Asian (6.6%) ancestry participants (Supplementary Tables 3 and 4).

### A multi-ancestry polygenic risk score with improved generalizability

The ancestry-specific GWAS meta-analyses identified 5,875, 945, 312, 702, and 79 independent variants associated with BMI (p-value < 1.0×10^−5^) in the European, African, Admixed American,East Asian, and South Asian ancestry groups, respectively. Variant weights for the corresponding ancestry-specific PRS are provided in Supplementary Table 5, and the weights obtained in the AoU model development dataset for combining these PRS into the MAPRS are provided in Supplementary Table 6.

In the AoU model evaluation dataset, the MAPRS outperformed individual ancestry-specific PRS and previously developed PRS based on European ancestry GWAS in capturing variance in measured BMI (Figure 2 and Supplementary Table 7). Specifically, the MAPRS achieved the highest Δ R^2^ both in the full model evaluation dataset (7.05%), and separately in the European (9.88%), African (2.60%), Admixed American (7.79%), East Asian (6.46%), and South Asian (7.23%) ancestry groups. The European ancestry-specific PRS developed in this study and a previously developed PRS using penalized regression (PGS001228) performed comparably to the MAPRS in the European ancestry group, but were outperformed by the MAPRS in all other ancestry groups (Figure 2 and Supplementary Table 7).

**Figure 2.**
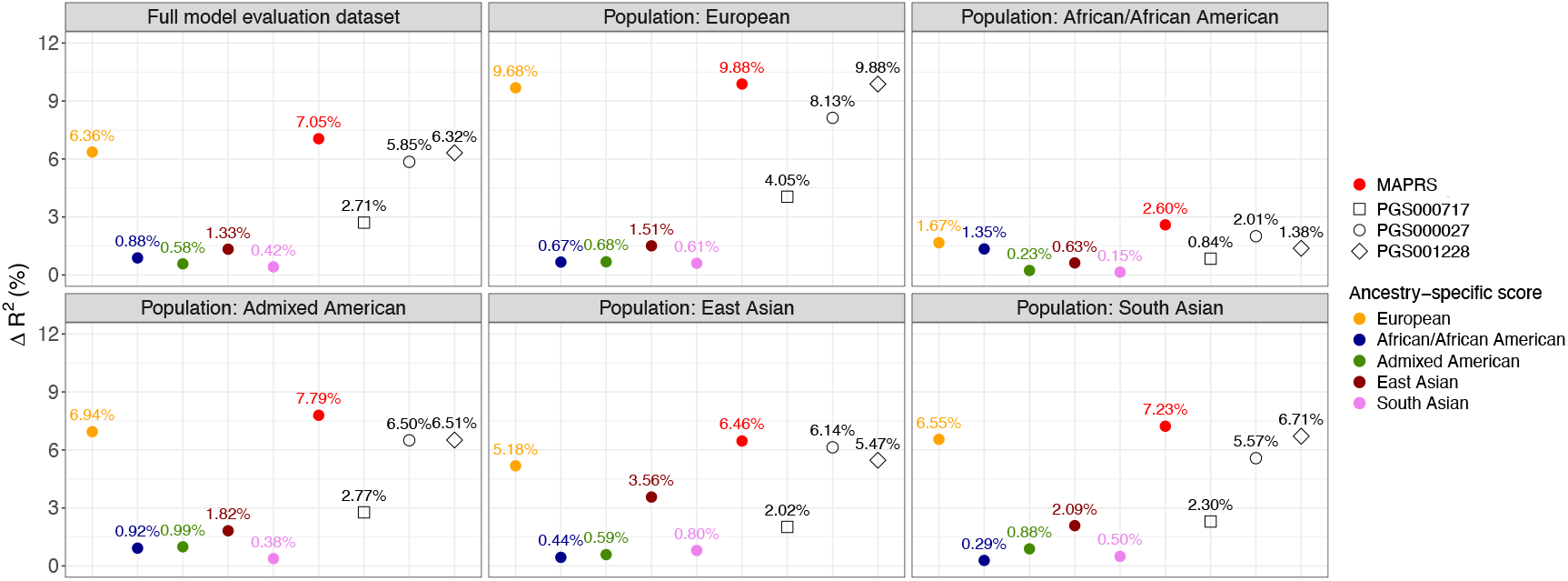
Performance of the MAPRS in predicting BMI in the AoU model evaluation dataset compared to ancestry-specific PRS. ΔR^2 represents the increase in the proportion of variance in BMI explained by adding the PRS to a baseline model. The MAPRS achieved the highest ΔR^2 both in the overall model evaluation dataset as well as within each genetic ancestry group. PGS000717, PGS000027, and PGS001228 are previously developed PRS based on European ancestry GWAS.

In the AoU test dataset, the MAPRS achieved similar Δ R^2^ in predicting baseline BMI (i.e., second-to-last measurement; 6.77%) and the most recent BMI (6.61%) (Supplementary Table 8). Within each ancestry group, the Δ R^2^ in the test dataset largely matched those obtained in the model evaluation dataset (Supplementary Table 8).

### Association between the multi-ancestry polygenic risk score and weight change outcomes

The MAPRS demonstrated significant associations with change in BMI as well as ≥10% and ≥5% TBW gain over 1.5-2.5 years in the AoU test dataset. A one SD increase in the MAPRS was associated with a 0.16 kg/m^2^ increase in BMI (standard error = 0.012 kg/m^2^; p-value = 2.2×10^−39^; Figure 3A and Supplementary Table 9), 1.27-fold increased odds of experiencing ≥10% TBW gain (95% CI: 1.24-1.31; p-value = 1.4×10^−55^; Figure 3B and Supplementary Table 10), and 1.15-fold increased odds of experiencing ≥5% TWB gain (95% CI: 1.13-1.18; p-value = 2.8×10^−39^; Figure 3B and Supplementary Table 11). These estimates were consistent between males and females, as indicated by overlapping sex-specific 95% confidence intervals (Supplementary Tables 9-11).

**Figure 3.**
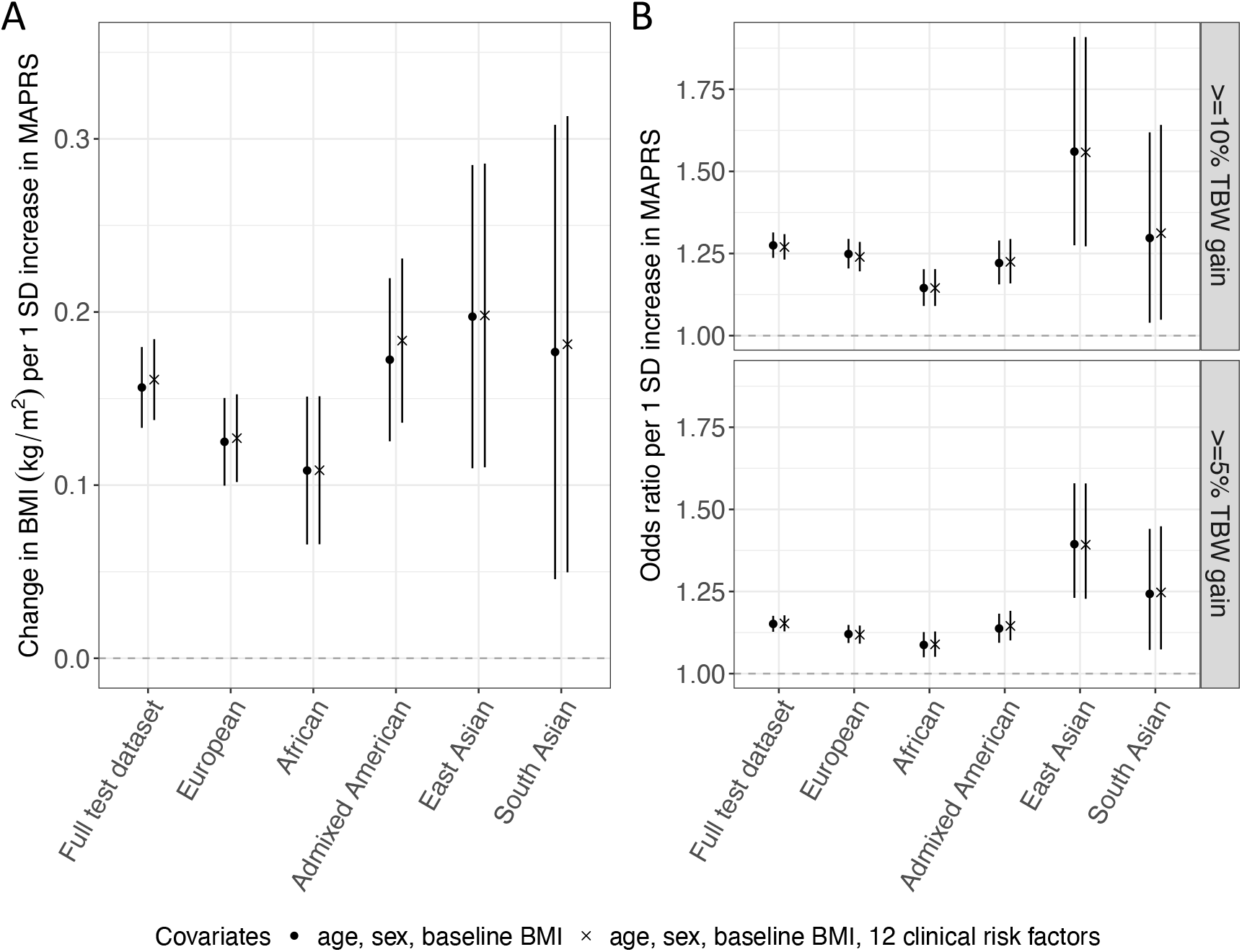
Association between the MAPRS and weight change outcomes in the AoU test dataset. (A) The association between the MAPRS and change in BMI was assessed using linear regression. (B) Associations between the MAPRS and ≥10% or ≥5% TBW gain were assessed using logistic regression. Each dot represents the estimated effect size per one standard deviation increase in the MAPRS, shown for the overall AoU test dataset and separately within each genetic ancestry group. Error bars indicate the 95% confidence intervals. All associations remained highly consistent after adjustment for clinical risk factors.

The magnitudes of associations were weakest among African ancestry participants and strongest among East Asian ancestry participants (Supplementary Tables 9-11). Specifically, within the African ancestry group, a one SD increase in the MAPRS was associated with 0.11 kg/m^2^ increase in BMI (standard error = 0.022 kg/m^2^; p-value = 6.5×10^−7^; Figure 3A), 1.14-fold increased odds of experiencing ≥10% TBW gain (95% CI: 1.09-1.20; p-value = 5.4×10^−8^; Figure 3B), and 1.09-fold increased odds of experiencing ≥5% TWB gain (95% CI: 1.05-1.13; p-value = 3.1×10^−6^; Figure 3B). In contrast, within the East Asian ancestry group, a one SD increase in the MAPRS was associated with 0.20 kg/m^2^ increase in BMI (standard error = 0.045 kg/m^2^; p-value = 1.1×10^−5^; Figure 3A), 1.56-fold increased odds of experiencing ≥10% TBW gain (95% CI: 1.28-1.91; p-value = 1.5×10^−5^; Figure 3B), and 1.39-fold increased odds of experiencing ≥5% TBW gain (95% CI: 1.23-1.58; p-value = 1.8×10^−7^; Figure 3B). Despite these discrepancies, the direction of association was consistent across all ancestry groups, and the ancestry-specific 95% confidence intervals largely overlapped with those in the overall test dataset (Figure 3A and B).

Notably, the MAPRS was significantly associated with each of the 12 investigated clinical risk factors in the AoU test dataset (Supplementary Figure 1 and Supplementary Table 12). For example, a one SD increase in the MAPRS was associated with 1.43-fold increased odds of developing type 2 diabetes (95% CI: 1.40-1.47; p-value = 1.7×10^−188^), with ancestry-specific odds ratios (ORs) ranging from 1.19 to 1.48 (Supplementary Figure 1 and Supplementary Table 12). However, adjusting for these clinical risk factors, either individually or simultaneously, did not attenuate the magnitude of association between the MAPRS and any of the weight change outcomes. Specifically, conditioned on all 12 clinical risk factors, a one SD increase in the MAPRS was still associated with a 0.16 kg/m^2^ increase in BMI (standard error = 0.012 kg/m^2^; p-value = 1.5×10^−41^; Figure 3A and Supplementary Table 9), 1.27-fold increased odds of experiencing ≥10% TBW gain (95% CI: 1.24-1.31; p-value = 1.5×10^−53^; Figure 3B and Supplementary Table 10), and 1.15-fold increased odds of experiencing ≥5% TBW gain (95% CI: 1.13-1.18; p-value = 1.1×10^−39^; Figure 3B and Supplementary Table 11). The ancestry-specific association estimates also remained highly consistent (Supplementary Tables 9-11).

### Comparison of genetic and clinical risk factors in association with weight change outcomes

The associations between clinical risk factors and weight change outcomes were inconsistent (Supplementary Tables 13-15). Among all 12 clinical risk factors, only obstructive sleep apnea was consistently associated with an increase in BMI (0.17 kg/m^2^; standard error = 0.031 kg/m^2^; p-value = 2.2×10^−8^; Figure 4A) and with increased odds of experiencing ≥10% TBW gain (OR = 1.30; 95% CI: 1.19-1.41; p-value = 2.7×10^−9^; Figure 4B) and ≥5% TBW gain (OR = 1.20; 95% CI: 1.14-1.28; p-value = 2.7×10^−9^; Figure 4C). In contrast, NAFLD was associated with a decrease in BMI (−0.15 kg/m^2^; standard error = 0.042 kg/m^2^; p-value = 2.8×10^−4^; Figure 4A) but with increased odds of experiencing ≥10% TBW gain (OR = 1.21; 95% CI: 1.08-1.36; p-value = 8.4×10^−4^; Figure 4B). Similarly, while type 2 diabetes was associated with a decrease in BMI (−0.20 kg/m^2^; standard error = 0.027 kg/m^2^; p-value = 3.0×10^−13^; Figure 4A), it was positively associated with odds of experiencing ≥10% or ≥5% TBW gain, although these positive associations did not withstand multiple testing correction (Figure 4B and C). Additionally, hypertension, GERD, CVD, osteoarthritis, chronic pain, depression, anxiety, and cholelithiasis were significantly associated with ≥10% TBW gain, but they were not associated with an increase in BMI (Figure 4A and B). Notably, the MAPRS was more strongly associated with all three weight change outcomes when comparing individuals above a MAPRS cutoff corresponding to the prevalence of each clinical risk factor versus the remainder of the population (Figure 4A-C).

**Figure 4.**
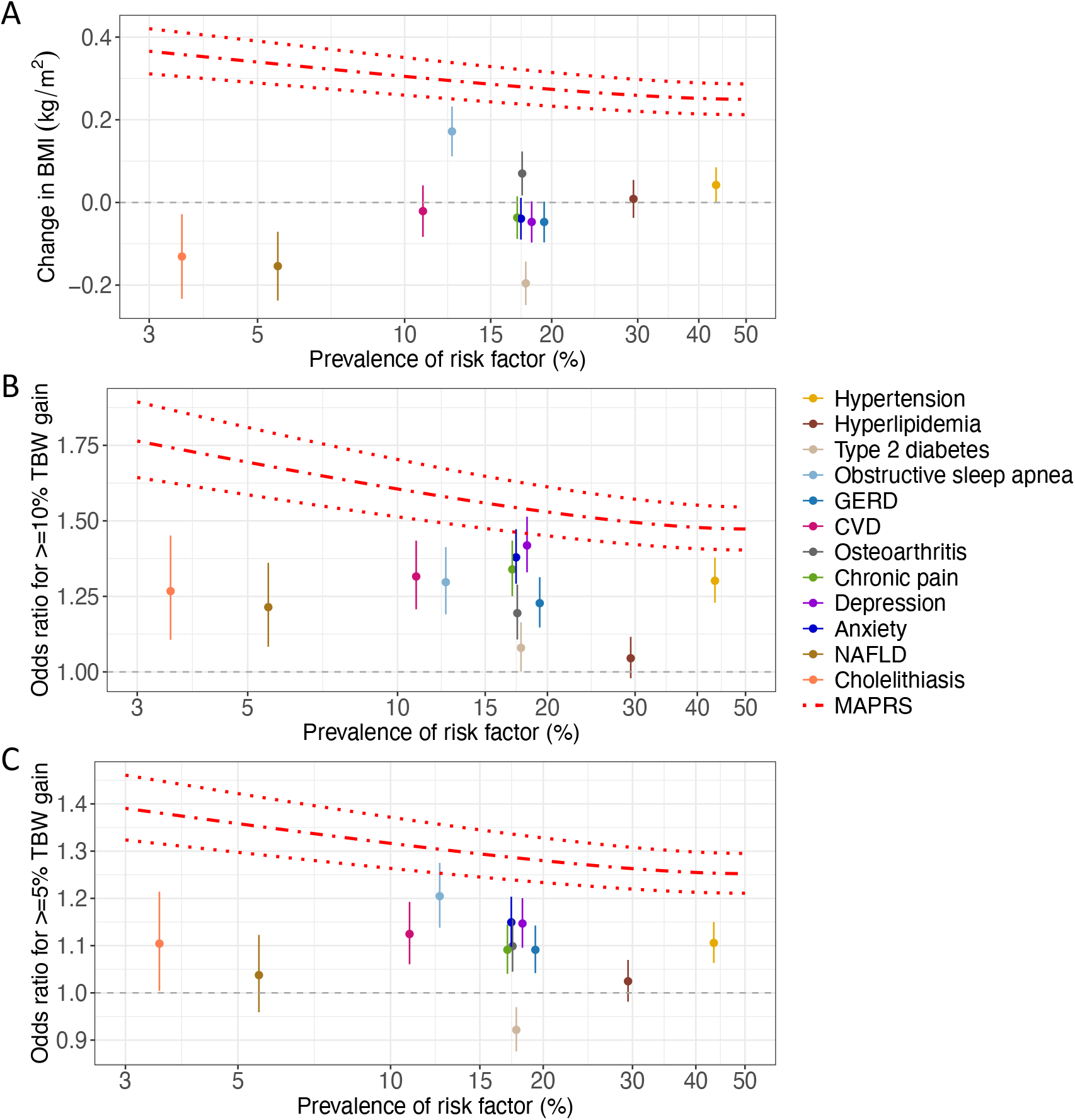
Comparison of MAPRS and clinical risk factor associations with weight change outcomes. Associations between each risk factor and (A) change in BMI, (B) 10% TBW gain, and (C) ≥5% TBW gain are shown in relation to their prevalence. Dots represent the estimated effect sizes in the overall AoU test dataset, with error bars indicating 95% confidence intervals. For the MAPRS, predicted effect sizes as a continuous risk factor are shown with respect to the proportion of individuals considered at risk, illustrated by the red dash-dotted line, with red dotted lines indicating the 95% confidence interval. X-axes are on a logarithmic scale.

## Discussion

In this study, we developed and validated an MAPRS for BMI, constructed by aggregating GWAS data across five genetic ancestries into a unified score. Compared to ancestry-specific PRS and previously developed PRS based on European ancestry GWAS, the MAPRS demonstrated improved generalizability, despite potential participation bias and resulting differences in demographic and health characteristics across genetic ancestry groups. Importantly, the MAPRS not only captured variation in BMI but also predicted prospective changes in BMI and the risk of significant weight gain over 1.5–2.5 years. In contrast, most of the 12 clinical risk factors we examined showed inconsistent or weak associations with weight change outcomes. These findings support the MAPRS as a robust and generalizable risk factor for obesity and weight gain.

The effectiveness of the MAPRS likely originates from the complex genetic architecture underlying BMI and weight regulation^8–11^. While large-scale GWAS are often limited in providing mechanistic insight, they capture a broad spectrum of biological pathways that may contribute to both baseline adiposity and changes in body weight over time. As a result, a PRS derived from GWAS of BMI may inadvertently capture a genetic propensity for weight gain. Notably, previous studies have shown that a PRS for BMI can predict weight trajectories from birth through adolescence and into early adulthood among individuals of European ancestry^12^. Our findings extend this understanding by demonstrating that genetic predisposition remains informative across diverse genetic ancestry groups and may be actionable even in middle-aged and older adults. Although future studies, including large-scale GWAS of weight change outcomes, are needed to elucidate the specific mechanisms driving these associations, these results highlight the enduring relevance of genetic predisposition in shaping weight trajectories.

Consistent with previous research showing overlap between genetic risk for obesity and its comorbidities^38^, the MAPRS was strongly associated with all 12 clinical risk factors. However, adjusting for these conditions did not attenuate the association between the MAPRS and weight change outcomes, suggesting that the clinical conditions are unlikely to mediate the observed genetic effects. Instead, these associations likely reflect the broad and pleiotropic effects of the genetic variants associated with BMI^8–11^. Importantly, lifestyle and environmental factors^39–41^, such as diet, physical activity, and socioeconomic status, can be associated with weight gain either independently or through interaction with genetic risk, but were not quantified in this study due to data limitations. Nevertheless, our findings suggest that the MAPRS may have clinical relevance in identifying individuals at risk of significant weight gain who could benefit from early lifestyle interventions^39,41^.

Interestingly, the MAPRS performed consistently whether the outcome was change in BMI, which accounts for height, or absolute weight gain (≥5% or ≥10%). In contrast, associations with clinical risk factors varied depending on the specific weight change outcome. This difference may reflect complex confounding and reverse causation in clinical conditions. For example, individuals with type 2 diabetes may experience weight loss due to weight management therapies or as a consequence of metabolic dysregulation associated with poor glycemic control^42^. Conversely, weight gain may occur in response to pharmacologic treatments, particularly insulin or sulfonylureas^43,44^. These opposing effects can obscure or weaken the overall association between clinical conditions and weight change. Moreover, whether height is accounted for or not can influence how these effects manifest, given the multifaceted and potentially bidirectional relationships between these clinical conditions, body size, and weight. This variability underscores the strength of the MAPRS, which captures a stable, lifelong genetic predisposition and is less susceptible to such biases.

This study has several limitations. First, while we demonstrated that the MAPRS is significantly associated with longitudinal weight change, we did not evaluate its performance in identifying high-risk individuals using more advanced classification or machine learning models. Future work could incorporate additional clinical and lifestyle factors into such models to further enhance predictive utility. Second, the GWAS data used to construct ancestry-specific PRS for non-European ancestry populations were based on smaller sample sizes, limiting their statistical power. Expanding GWAS in underrepresented populations remains crucial for building equitable and generalizable MAPRS. Third, to maximize sample size, the AoU model development, evaluation, and test datasets were defined based on the availability of BMI measurements. This may have introduced selection bias, as suggested by differences in demographic characteristics across these datasets. However, the consistent performance of the MAPRS across datasets suggests that any such bias likely had minimal impact on the association estimates. Future studies using additional cohorts with increased sample sizes may benefit from stratified sampling or harmonized study designs to further mitigate this issue. Additionally, our PRS construction strategy favored simplicity and cost-efficiency by using a clumping-and-thresholding approach with fewer variants. Future work may explore more advanced methods, such as XPASS^45^, PRS-CSx^46^, or X-WING^47^, to potentially improve predictive accuracy, albeit with greater computational and clinical implementation complexity. Lastly, our longitudinal analysis was limited to a follow-up period of 1.5–2.5 years. Extending the observation period in future studies will help clarify the extent to which genetic risk predicts long-term weight trajectories.

## Conclusions

In conclusion, the MAPRS developed in this study is associated not only with current BMI but also with longitudinal weight change, offering a robust measure of genetic predisposition to future weight gain. The improved generalizability across diverse genetic ancestries and consistency across different outcome definitions of the MAPRS highlight its promise as a tool for genetic risk stratification and for informing early, targeted interventions in preventive healthcare.

## Acknowledgements

We gratefully acknowledge All of Us participants for their contributions, without whom this research would not have been possible. We also thank the National Institutes of Health’s All of Us Research Program for making available the participant data examined in this study. T.L. has been supported by start-up funding from the Office of the Vice Chancellor for Research and Graduate Education, School of Medicine and Public Health, and Department of Population Health Sciences at the University of Wisconsin-Madison.

## Data Availability

Access to the All of Us Research Program is governed by the program’s data access policies and is available to registered researchers in accordance with those guidelines. GWAS summary statistics are available at the Figshare repository: https://figshare.com/s/63add42ab71725a30cb7. Variant-level weights used in the polygenic risk scores are available in Supplementary Table 5.

## Author Contributions

T.L. conceptualized the study. T.L. and W.Z. designed the methodology. L.M.F., L.N.S., K.V.L., and B.M.H. acquired the data. T.L. performed data analysis with contributions from all authors. T.L. wrote the initial draft of the manuscript. All authors interpreted the results, revised the manuscript critically, and approved the final version of the manuscript.

---

**Supplementary Figure S1.**
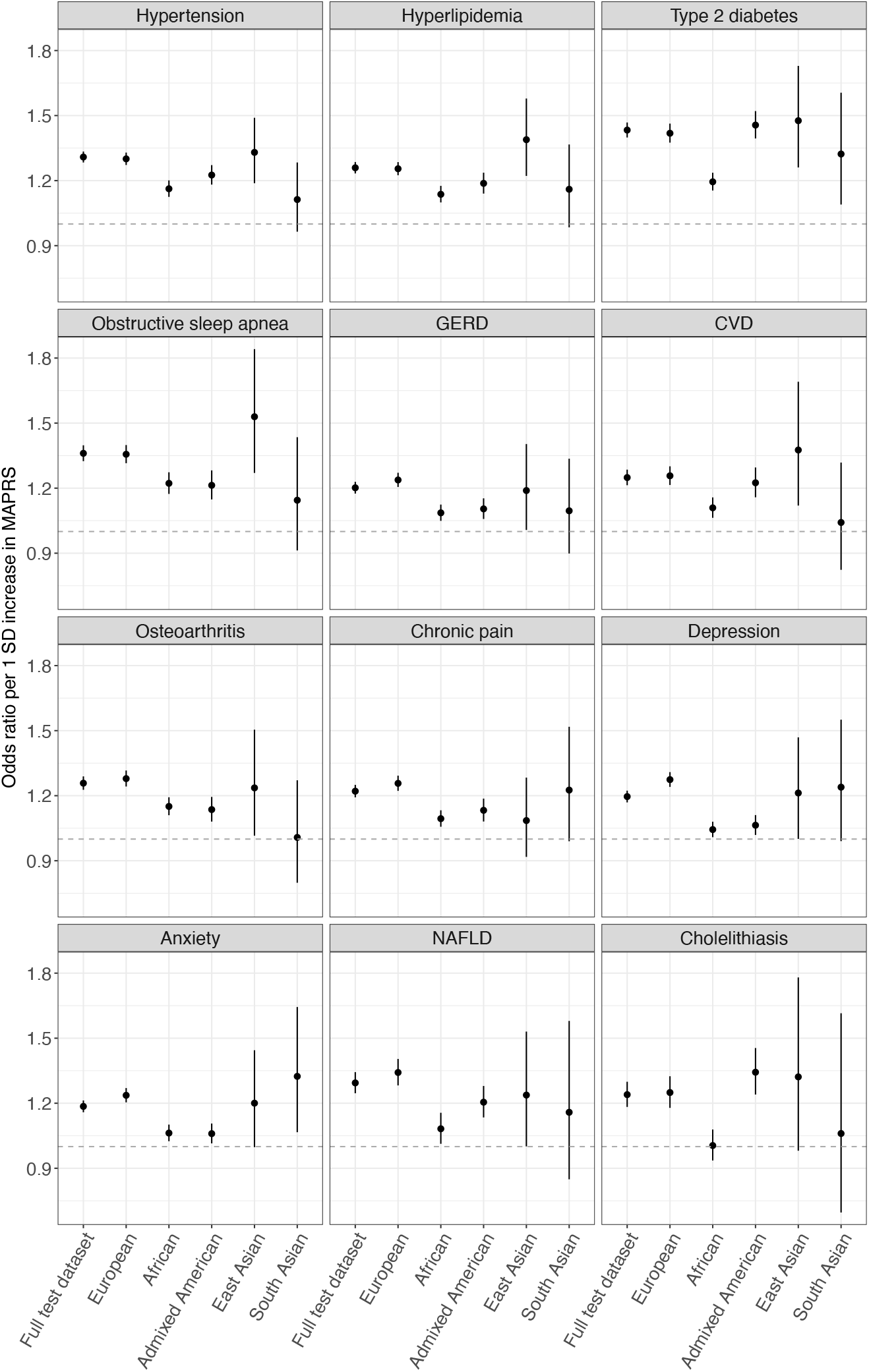
Associations between the MAPRS and 12 clinical risk factors. Each dot represents the odds ratio for having the indicated clinical risk factor per one standard deviation increase in the MAPRS, estimated in the overall AoU test dataset and separately within each genetic ancestry group. Error bars indicate the 95% confidence intervals.

